# Vascular injury markers associated with cognitive impairment in people with HIV on suppressive antiretroviral therapy

**DOI:** 10.1101/2023.07.23.23293053

**Authors:** Debjani Guha, Vikas Misra, Jun Yin, Miki Horiguchi, Hajime Uno, Dana Gabuzda

## Abstract

**Objective:** Human immunodeficiency virus (HIV)-associated neurocognitive disorders (HAND) remain prevalent despite viral suppression on antiretroviral therapy (ART). Vascular disease contributes to HAND, but peripheral markers that distinguish vascular cognitive impairment (VCI) from HIV-related etiologies remain unclear.

**Design:** Cross-sectional study of vascular injury, inflammation, and central nervous system (CNS) injury markers in relation to HAND.

**Methods:** Vascular injury (VCAM-1, ICAM-1, CRP), inflammation (IFN-γ, IL-1β, IL-6, IL-8, IL-15, IP-10, MCP-1, VEGF-A), and CNS injury (NFL, total Tau, GFAP, YKL-40) markers were measured in plasma and CSF from 248 individuals (143 HIV+ on suppressive ART and 105 HIV-controls).

**Results:** Median age was 53 years, median CD4 count, and duration of HIV infection were 505 cells/µl and 16 years, respectively. Vascular injury, inflammation, and CNS injury markers were increased in HIV+ compared with HIV-individuals (p<0.05). HAND was associated with increased plasma VCAM-1, ICAM-1, and YKL-40 (p<0.01) and vascular disease (p=0.004). In contrast, inflammation markers had no significant association with HAND. Vascular injury markers were associated with lower neurocognitive T scores in age-adjusted models (p<0.01). Furthermore, plasma VCAM-1 correlated with NFL (r=0.29, p=0.003). Biomarker clustering separated HAND into three clusters: two clusters with high prevalence of vascular disease, elevated VCAM-1 and NFL, and distinctive inflammation profiles (CRP/ICAM-1/YKL-40 or IL-6/IL-8/IL-15/MCP-1), and one cluster with no distinctive biomarker elevations.

**Conclusions:** Vascular injury markers are more closely related to HAND and CNS injury in PWH on suppressive ART than inflammation markers and may help to distinguish relative contributions of VCI to HAND.

## INTRODUCTION

Combination antiretroviral therapy (ART) has reduced HIV-related morbidity and mortality, but mild forms of HIV-associated neurocognitive disorders (HAND), consisting of asymptomatic neurocognitive impairment (ANI) and mild neurocognitive disorder (MND), remain prevalent in people with HIV (PWH) [1–3]. The mechanisms underlying HAND in PWH on suppressive ART are not well characterized [3, 4]. Although viral reservoirs in brain can impact neuroinflammation and cognitive function [3, 5, 6], other factors including systemic immune activation/inflammation, oxidative stress, metabolic abnormalities, blood-brain barrier (BBB) dysfunction, and ART drug neurotoxicity contribute to mechanisms involved in HAND in ART-treated PWH [4, 7–9]. Comorbidities including vascular disease and substance abuse also impact the development of HAND [3, 4, 10, 11]. The identification of biomarkers associated with cognitive impairment and central nervous system (CNS) injury due to HIV-related vs. non-HIV-related etiologies is important for understanding biologically defined subtypes (biotypes) of HAND and development of tailored interventions [2, 12, 13].

Vascular disease is more common in PWH compared with the general population [14–16]. The increased prevalence of atherosclerosis, coronary artery disease, and cerebrovascular disease reflects HIV-related factors including immune activation/inflammation, metabolic abnormalities, and effects of some ART drugs, as well as high rates of smoking and substance use [14–18]. Due to earlier onset and progression of vascular disease, PWH are also at increased risk for vascular cognitive impairment (VCI) at younger ages compared with the general population [10, 19]. Given evidence that VCI contributes to cognitive impairment in PWH on ART [17, 20, 21], vascular disease has emerged as a significant contributing factor to HAND in the current ART era [10, 19, 22–25]. Accordingly, biomarkers that can distinguish VCI from other causes of cognitive impairment will be useful for diagnosis and clinical management.

The relationship between markers of vascular injury and HAND in virally suppressed PWH is unclear. Vascular injury markers (e.g., vascular cell adhesion molecule [VCAM-1], intercellular adhesion molecule [ICAM-1], selectins) are increased in PWH on ART [14, 18, 25–28]. Increased expression of endothelial activation markers such as VCAM-1 and ICAM-1 is associated with systemic and arterial inflammation in PWH on ART [18, 27, 29]. Consistent with these findings, inflammation markers including interleukin (IL)-6, C-reactive protein (CRP), interferon gamma-induced protein (IP)-10, monocyte chemoattractant protein (MCP)-1, and vascular endothelial growth factor (VEGF)-A are associated with vascular disease in ART-treated PWH [18, 30–32]. Plasma and/or cerebrospinal fluid (CSF) neurofilament light chain (NFL), total and phosphorylated Tau, glial fibrillary acidic protein (GFAP), and YKL-40 (also known as Chitinase 3-like 1, CHI3L1) are biomarkers of CNS injury and glial activation in PWH [12, 33–39]. Here, we evaluated the relationship between vascular injury markers and HAND by assessing plasma and CSF markers of vascular injury (ICAM-1, VCAM-1, CRP), inflammation (IL-1β, IFN-γ, IL-6, IL-8, IL-15, IP-10, MCP-1, VEGF-A), and CNS injury and glial activation (NFL, total Tau, GFAP, YKL-40) in relation to cognitive impairment in PWH on suppressive ART.

## METHODS

### Study participants

Plasma and CSF samples from 248 eligible individuals (n=143 PWH on ART and n=105 HIV-controls) were collected during 2006–2016 (Supplementary Digital Content 1). HIV-positive samples were from the National NeuroAIDS Tissue Consortium (NNTC) [40] and CNS HIV Anti-Retroviral Therapy Effects Research (CHARTER) study [41]. All individuals were enrolled with written informed consent and institutional review board (IRB) approval at each study site.

All participants were over age 30 years and PWH were taking 3 or more ART drugs for at least 1 year with current viral suppression in plasma (<200 HIV RNA copies/ml). Eight individuals with plasma viral loads slightly above 200 copies/ml were also included (median viral load 357 copies/ml [IQR 323–425]). PWH with HIV-associated dementia (HAD) or neuropsychological impairment due to other causes (NPI-O) were excluded because HAD diagnoses are rare in the current ART era and NPI-O reflects confounding diagnoses. Plasma and CSF samples from HIV-individuals without a diagnosed neurological disease (from Bioreclamation LLC, Westbury, New York) were group-matched for age, gender, and race.

### Assessment of cognitive impairment and medical comorbidities

PWH were administered a comprehensive neuropsychological test battery designed to assess seven neurocognitive domains. Demographically corrected global neurocognitive T scores were generated from individual test T scores as described [42]. T scores correlate negatively with severity of cognitive impairment, with values below 40 (corresponding to 1 standard deviation of 10 below a normalized mean of 50) signifying cognitive impairment. HAND clinical diagnoses were determined using established criteria [43] based on neurocognitive testing and neurological evaluation. PWH were classified as cognitively impaired if they had a clinical HAND diagnosis of ANI or MND, corresponding to mild cognitive impairment with no interference or mild interference with everyday functioning, respectively. A history of medical comorbidities (e.g., hypertension, hyperlipidemia, diabetes, coronary artery disease, myocardial infarction, cerebrovascular disease, ischemic stroke, lacunes) was obtained from self-report, medical records, and review of medications and lab values as described [44]. Traditional cardiovascular risk factors were defined based on ≥2 visits with self-reported use of medications for these conditions or laboratory test values as follows: hypertension, systolic blood pressure >140 or diastolic blood pressure >90 mm Hg; hyperlipidemia, total cholesterol >240 mg/dl; diabetes, hemoglobin A1C ≥6.5%.

### Meso Scale Discovery assays

Plasma and CSF samples were centrifuged at 1200 rpm for 5 minutes at 4°C to remove cells, and supernatants were aliquoted and stored at -80°C. Vascular injury (ICAM-1, VCAM-1, CRP), inflammation (IFN-γ, IL-1β, IL-6, IL-8, IL-15, IP-10, MCP-1, VEGF-A), CNS injury and glial activation (NFL, total Tau, GFAP, YKL-40) markers were measured in plasma and CSF using the Meso Scale Discovery (MSD) platform (Rockville, MD). Vascular injury markers were measured using V-Plex vascular injury panel 2, inflammation markers using a U-Plex custom panel, and CNS injury and glial activation markers using R-Plex custom panels according to manufacturer’s protocols. Plates were read on Meso Sector S 600 imager and data analyzed using MSD discovery workbench 4.0 software.

### Measurement of CSF/plasma albumin ratio (Qalb)

Albumin concentrations in paired plasma and CSF samples were measured using the BCG albumin assay kit (Thermo Fisher Scientific). Absorbance was measured at 570 nm, concentrations calculated using standard curves, and CSF/plasma albumin ratio (Qalb) calculated as an indirect indicator of BBB permeability.

### Statistical analysis

Demographics, clinical covariates, and biomarker levels were compared between two groups of interest using the chi-square test for categorical variables and Mann-Whitney U test for continuous variables. Relationships between continuous variables were analyzed by Spearman’s rank correlation. Associations between plasma vascular or CNS injury markers and global neurocognitive T scores were examined in linear and logistic regression models adjusted for age with T scores as a continuous or binary (<40 vs. ≥40) dependent variable, respectively, and log_10_-transformed plasma biomarkers as predictors. A two-sided p-value <0.05 was considered statistically significant. These analyses were performed using GraphPad Prism, version 9.0 (GraphPad Software, Inc., La Jolla, California). For biomarker-driven clustering of HAND (n=71 after omitting 3 cases with missing values), the following R packages were used (R version 4.2.0, R Foundation for Statistical Computing, Vienna, Austria): principal component analysis (PCA) was performed with 15 biomarkers using the FactoMineR package, K-means clustering was performed using the stats package, and clusters were visualized using the factoextra package.

## RESULTS

### Study cohort

Demographic and clinical characteristics of the study population are shown in Table 1 and overview of the study design is shown in Supplementary Digital Content 1. The cohort consisted of 143 PWH (median age 53 years [IQR 47–58 years], 85% male, 71% white, median duration of HIV infection 16 years, median CD4 and nadir CD4 counts 505 and 88 cells/µl, respectively) and 105 HIV-controls matched for demographics. PWH were on ART with 94% and 96% having plasma and CSF viral load (VL) <200 copies/ml and <50 copies/ml, respectively. Among PWH, 52% were diagnosed with mild forms of HAND (59.4% ANI, 40.6% MND). Cardiovascular and cerebrovascular disease (CVD) (e.g., coronary artery disease, myocardial infarction, ischemic stroke, silent brain infarcts, lacunes; hereafter termed vascular disease) was more prevalent among PWH with HAND (26%), particularly MND (40%), compared with no HAND (7%) (p=0.004 and p=0.0006, respectively) (Table 1 and Supplementary Digital Content 2). Prevalence of 2-3 cardiovascular risk factors was also more common in HAND (30%) or MND (40%) vs. no HAND (17%) (p=0.12 and p=0.005, respectively). PWH with vascular disease were slightly older (median age 56 vs. 51 years; p=0.02), with higher prevalence of tobacco smoking (75% vs. 42%; p=0.003), hypertension (63% vs. 44%; p=0.09), hyperlipidemia (38% vs. 22%; p=0.10), and HAND (79% vs. 46%; p=0.003) and lower neurocognitive T scores (median 42 vs. 49; p=0.002) compared to those without vascular disease (Supplementary Digital Content 2).

**Table 1.** Demographic and clinical characteristics of study participants.

|  | HIV-<br>(n=105) | HIV+<br>(n=143) | No HAND<br>(n=69) | HAND<br>(n=74) | P-value |
| --- | --- | --- | --- | --- | --- |
| Age (years) | 54 (47 - 61) | 53 (47 - 58) | 53 (47 - 58) | 52 (47 - 58) | 0.98 |
| Male gender (n, %) | 74 (70) | 121 (85) | 62 (90) | 58 (78) | 0.07 |
| Race (n, %)* |  |  |  |  | 0.98 |
| Black | 22 (27) | 36 (25) | 18 (26) | 18 (24) |  |
| White | 58 (72) | 101 (71) | 48 (70) | 53 (72) |  |
| Other | 1 (1) | 6 (4) | 3 (4) | 3 (4) |  |
| Current smoking (n, %) |  | 68 (48) | 27 (39) | 41 (55) | 0.07 |
| Current alcohol use (n, %) |  | 67 (47) | 34 (49) | 34 (46) | 0.74 |
| BMI (kg/m<sup>2</sup>) |  | 26 (22 - 30) | 25 (22 - 28) | 27 (22 - 33) | 0.12 |
| Diabetes (n, %) |  | 21 (15) | 8 (12) | 13 (18) | 0.35 |
| Hypertension (n, %) |  | 67 (47) | 32 (46) | 35 (47) | 1.00 |
| Hyperlipidemia (n, %) |  | 35 (25) | 14 (20) | 21 (28.3) | 0.33 |
| 2-3 Cardiovascular risk factors (n, %)* |  | 35 (25) | 12 (17) | 22 (30) | 0.12 |
| Cerebrovascular disease/CVD (n, %) |  | 24 (17) | 5 (7) | 19 (26) | **0.004** |
| Neurocognitive T scores |  | 48 (42 - 53) | 52 (49 - 57) | 42 (38 - 47) | **<0.0001** |
| Global clinical rating |  | 4 (3 - 5) | 3 (2 - 4) | 5 (4 - 6) | **<0.0001** |
| CSF protein |  | 37 (31 - 47) | 41 (32 - 50) | 36 (30 - 45) | 0.07 |
| CSF WBC (cells/μl) | | 2 (1 - 3) | 2 (1 - 3) | 1 (1 - 4) | 0.77 |
| Duration of HIV infection (years) |  | 16 (11 - 21) | 16 (11 - 21) | 16 (11 - 21) | 0.90 |
| Plasma viral load <200 copies/ml |  | 134 (94) | 65 (94) | 69 (95) | 1.00 |
| CSF viral load <50 copies/ml<sup>#</sup> |  | 122 (96) | 60 (97) | 61 (94) | 0.68 |
| CD4 count (cells/μl) | | 505 (328 - 734) | 504 (339 - 827) | 506 (291 - 681) | 0.41 |
| Nadir CD4 count (cells/μl) | | 88 (17 - 211) | 88 (13 - 244) | 91 (21 - 186) | 0.84 |
| ART use (n, %) |  |  |  |  | 0.69 |
| Protease inhibitors |  | 84 (59) | 41 (59) | 43 (58) |  |
| Integrase inhibitors |  | 27 (19) | 12 (17) | 15 (20) |  |
| Duration of ART (years) |  | 10 (6 - 14) | 10 (6 - 14) | 9 (5 - 14) | 0.67 |

Median (interquartile range) are shown unless otherwise indicated. P-values for two-group comparisons between HAND vs.no HAND were calculated using chi-square test for categorical variables and Mann-Whitney U test for continuous variables; bold font denotes p <0.05. HAND diagnoses were asymptomatic neurocognitive impairment (ANI, n= 44) or mild neurocognitive disorder (MND, n= 30). \*Race data not available for 24 HIV- individuals. <sup>#</sup>CSF viral load data not available for 16 HIV+ individuals. <sup>†</sup>2-3 cardiovascular risk factors included diabetes, hyperlipidemia, and hypertension. ART, antiretroviral therapy; BMI, body mass index; CVD, cardiovascular disease; HAND, HIV-associated neurocognitive disorders; WBC, white blood cells.

### Association of vascular injury markers with HAND

To evaluate relationships of vascular injury markers to HAND, we compared levels of plasma ICAM-1, VCAM-1, and CRP between groups by HIV status, HAND, and vascular disease (Figure 1A). Plasma ICAM-1 and CRP, but not VCAM-1, were increased in PWH compared with HIV-controls (p=0.004, p=0.0005, and p=0.11, respectively). Among PWH, ICAM-1 and VCAM-1 increased (p=0.01), and CRP showed an increasing trend (p=0.08) in HAND vs. no HAND. We further examined the association of vascular injury markers with ANI and MND (Supplementary Digital Content 3). VCAM-1 was higher in PWH with MND compared with ANI (p=0.04) or no HAND (p=0.001), while ICAM-1 was higher in ANI and showed an increasing trend in MND (p=0.007 and p=0.18, respectively). Plasma VCAM-1, but not ICAM-1 or CRP, was higher in PWH with vs. without vascular disease (p=0.049) (Figure 1A). Neither HIV infection nor HAND were associated with increased levels of these markers in CSF (Figure 1B).

**Figure 1.**
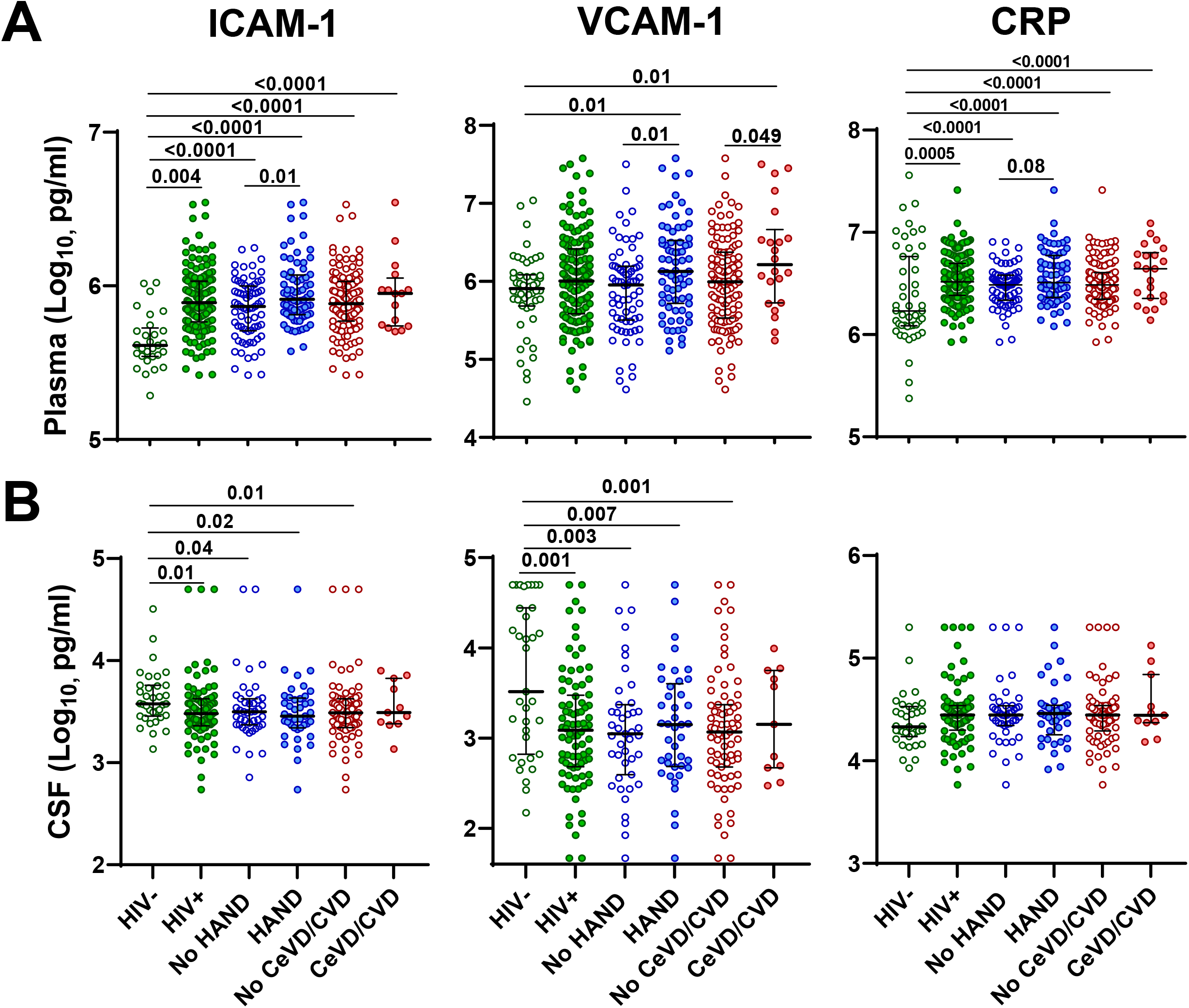
HIV infection and HAND are associated with increased plasma vascular injury markers. (A) Association of plasma ICAM-1, VCAM-1, and CRP with HIV infection, HAND, and vascular disease. (B) Association of CSF ICAM-1, VCAM-1, and CRP with HIV infection, HAND, and vascular disease. Medians and IQRs are indicated as horizontal and vertical lines, respectively. Statistical significance was calculated using Mann–Whitney U test; significant differences (p<0.05) are indicated.

### CNS injury markers in relation to HAND and vascular disease

Next, we examined associations of CNS injury and glial activation markers with HIV, HAND, and vascular disease (Figure 2). HIV infection was associated with increased plasma NFL and GFAP (p=0.004 and p=0.005, respectively), and increased CSF total Tau and GFAP (p=0.03 and p<0.0001, respectively). HAND was associated with increased plasma and CSF YKL-40 (p=0.004 and p=0.003, respectively), and trend toward increased CSF Tau (p=0.07) compared with no HAND (Figure 2A). Although we did not detect association of plasma NFL with HAND, we observed an increasing trend of plasma NFL (p=0.06) in MND vs. ANI (Supplementary Digital Content 3) and increased plasma NFL and GFAP in PWH with vs. without vascular disease (p=0.004 and p=0.04, respectively) (Figure 2A). There was no significant association of HAND with CSF/plasma albumin ratio (Qalb) (median 4.9 vs. 3.7 in HAND vs. no HAND, respectively; p=0.68).

**Figure 2.**
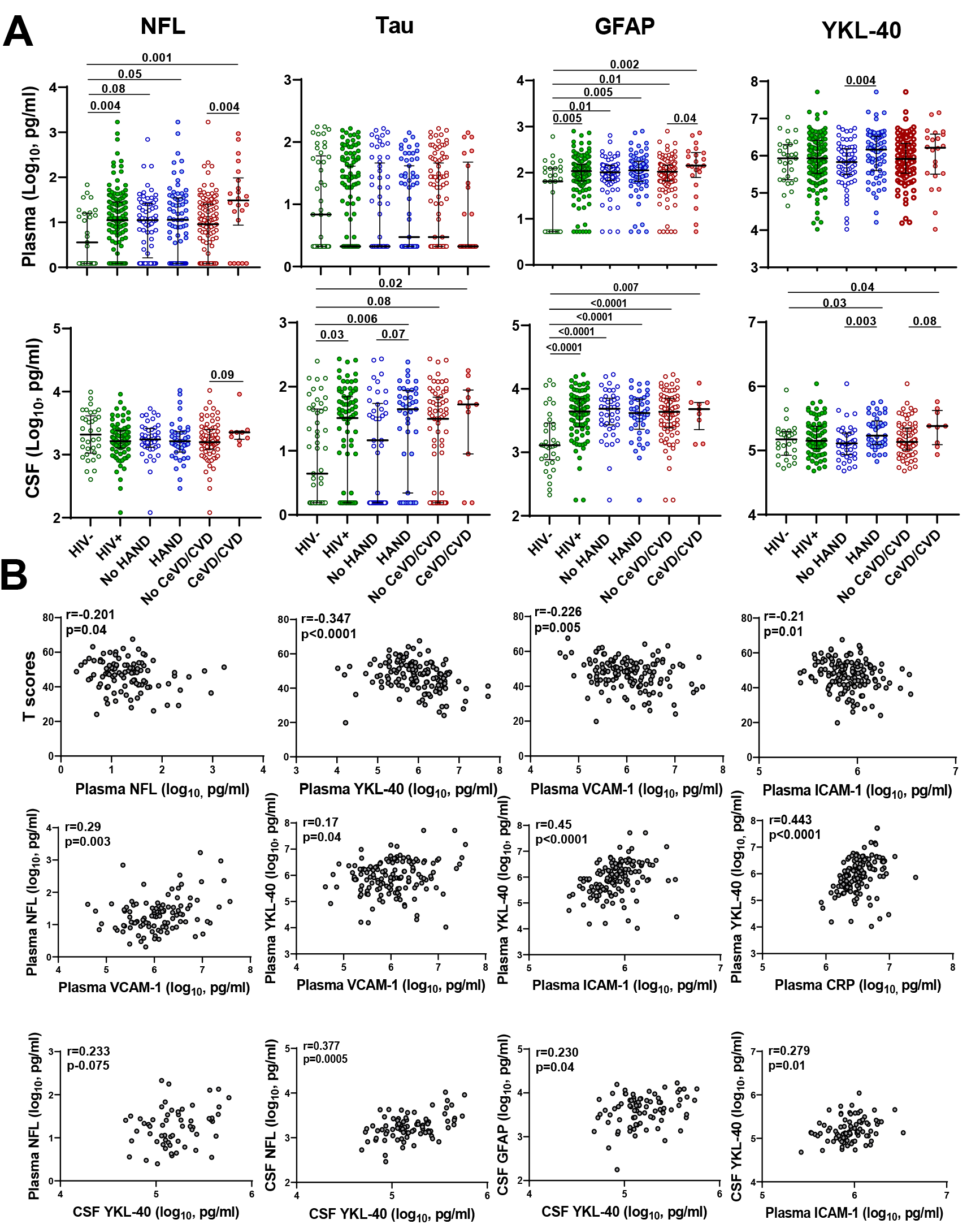
Vascular injury markers are associated with lower neurocognitive T scores and correlate with CNS injury markers. (A) Association of plasma (top) and CSF (bottom) NFL, total Tau, GFAP, and YKL-40 with HIV infection, HAND, and vascular disease. Medians and IQRs are indicated as horizontal and vertical lines, respectively. Statistical significance was calculated using Mann–Whitney U test; significant differences (p<0.05) are indicated. (B) Plasma NFL, YKL-40, ICAM-1, and VCAM-1 levels correlate negatively with global neurocognitive T scores, plasma VCAM-1 correlates positively with plasma NFL, and plasma VCAM-1, ICAM-1, and CRP correlate positively with plasma YKL-40 in PWH (middle panels). Inter-relationship of plasma and CSF CNS injury and glial activation markers with plasma vascular injury markers are shown in the bottom panels. Relationships between continuous variables were analyzed by Spearman’s rank correlation (significant correlations p<0.05).

### Vascular injury markers correlate with lower neurocognitive test scores

Next, we evaluated relationships between vascular or CNS injury markers and global neurocognitive T scores, which reflect severity of HAND [9]. Plasma NFL, YKL-40, VCAM-1, and ICAM-1 levels correlated negatively with global T scores (Figure 2B), while no significant correlations were detected between GFAP or CSF biomarkers and global T scores. In linear regression models adjusted for age (Table 2), higher plasma NFL (β=-4.06, p=0.019), YKL-40 (β=-3.57, p=0.001), VCAM-1 (β=-3.57, p=0.003), ICAM-1 (β=-9.05, p=0.008), and CRP (β=-8.37, p=0.007), but not GFAP (p=0.2) were associated with lower global T scores. The models were not adjusted for gender or race because global T scores were not significantly different between males vs. females (p=0.97) or white vs. non-white race (p=0.75). We further assessed association of CNS and vascular injury markers in logistic regression models with cognitive impairment defined as global T scores <40 vs. ≥40, corresponding to the impaired vs. unimpaired range, respectively (Supplementary Digital Content 4). Plasma NFL (β=1.057, p=0.03), YKL-40 (β=1.23, p=0.004), VCAM-1 (β=1.22, p=0.003), and ICAM-1 (β=2.38, p=0.03) but not GFAP (p=0.09) were associated with global T scores <40 in these models.

**Table 2.** Association of plasma NFL, GFAP, YKL-40, ICAM-1, VCAM-1, and CRP with global neurocognitive T scores in linear regression analysis adjusted for age.

| Variable | $\beta$ | SE | 95% CI | P-value |
| --- | --- | --- | --- | --- |
| **Intercept** | 50.21 | 2.38 | 45.5 to 54.9 | <0.0001 |
| **Age** | 0.16 | 0.11 | -0.07 to 0.38 | 0.16 |
| **NFL** | -4.06 | 1.70 | -7.42 to -0.70 | 0.019 |
| **Intercept** | 53.12 | 4.52 | 44.2 to 62.1 | <0.0001 |
| **Age** | -0.011 | 0.086 | -0.18 to 0.16 | 0.90 |
| **GFAP** | -2.88 | 2.26 | -7.34 to 1.58 | 0.20 |
| **Intercept** | 69.16 | 6.61 | 56.1 to 82.2 | <0.0001 |
| **Age** | -0.061 | 0.079 | -0.22 to 0.096 | 0.44 |
| **YKL-40** | -3.57 | 1.084 | -5.72 to -1.43 | 0.001 |
| **Intercept** | 101.60 | 20.27 | 61.5 to 141.7 | <0.0001 |
| **Age** | -0.079 | 0.081 | -0.24 to 0.081 | 0.33 |
| **ICAM-1** | -9.05 | 3.40 | -15.77 to -2.33 | 0.008 |
| **Intercept** | 68.93 | 7.18 | 54.73 to 83.13 | <0.0001 |
| **Age** | -0.018 | 0.080 | -0.17 to 0.14 | 0.82 |
| **VCAM-1** | -3.57 | 1.19 | -5.92 to -1.21 | 0.003 |
| **Intercept** | 102.60 | 20.04 | 63.0 to 142.2 | <0.0001 |
| **Age** | -0.067 | 0.080 | -0.23 to 0.091 | 0.40 |
| **CRP** | -8.37 | 3.052 | -14.4 to -2.34 | 0.007 |

Multivariable linear regression models were fit for global T scores in PWH as the dependent variable adjusting for age. Independent variables (plasma NFL, GFAP, YKL-40, ICAM-1, VCAM-1, CRP) were log<sub>10</sub> transformed. SE, standard error; CI, confidence interval.

### Vascular injury markers correlate with CNS injury and glial activation markers

To assess inter-relationships between vascular and CNS injury markers in PWH, we performed correlation analyses. Plasma VCAM-1 correlated with plasma NFL and YKL-40 (r=0.29, p=0.003; r=0.17, p=0.04, respectively), while plasma ICAM-1 and CRP correlated with plasma YKL-40 (r=0.45, p<0.0001 and r=0.443, p<0.0001, respectively) and CSF YKL-40 (r=0.279, p=0.01 and r=0.288, p=0.008) (Figure 2B). CSF YKL-40 correlated with plasma NFL, CSF NFL, and CSF GFAP (r=0.233, p=0.075; r=0.377, p=0.0005 and r=0.230, p=0.04, respectively). Plasma ICAM-1, CRP, and NFL correlated with CSF YKL-40, but showed no significant association with increased Qalb (Supplementary Digital Content 5).

### Inflammation markers in relation to HAND and vascular disease

Given that inflammation is a contributing factor to development of HAND [4, 8, 37, 45], we examined inflammation marker levels by HIV status, HAND, and vascular disease. Plasma IL-1β, IL-8, IP-10, MCP-1, and VEGF-A were higher (p=0.02, p=0.002, p<0.0001, p=0.003, and p<0.0001, respectively) in PWH compared with HIV-controls (Figure 3A). Plasma IL-6 was increased in HAND, ANI, or MND, vs. no HAND (p<0.0001, p=0.01, and p<0.0001, respectively), while IL-15, IFN-γ, and VEGF-A were increased in MND compared with no HAND (p=0.01, p=0.02, and p=0.04, respectively) (Figure 3A and Supplementary Digital Content 3). Plasma IL-6 and MCP-1 were increased (p=0.03 and p=0.01, respectively) and IL-15 showed an increasing trend (p=0.05) (Figure 3A) in PWH with vs. without vascular disease. Among CSF markers, only IP-10 showed a significant difference by HIV status (p=0.02), and none were significantly associated with HAND (Supplementary Digital Content 6).

**Figure 3.**
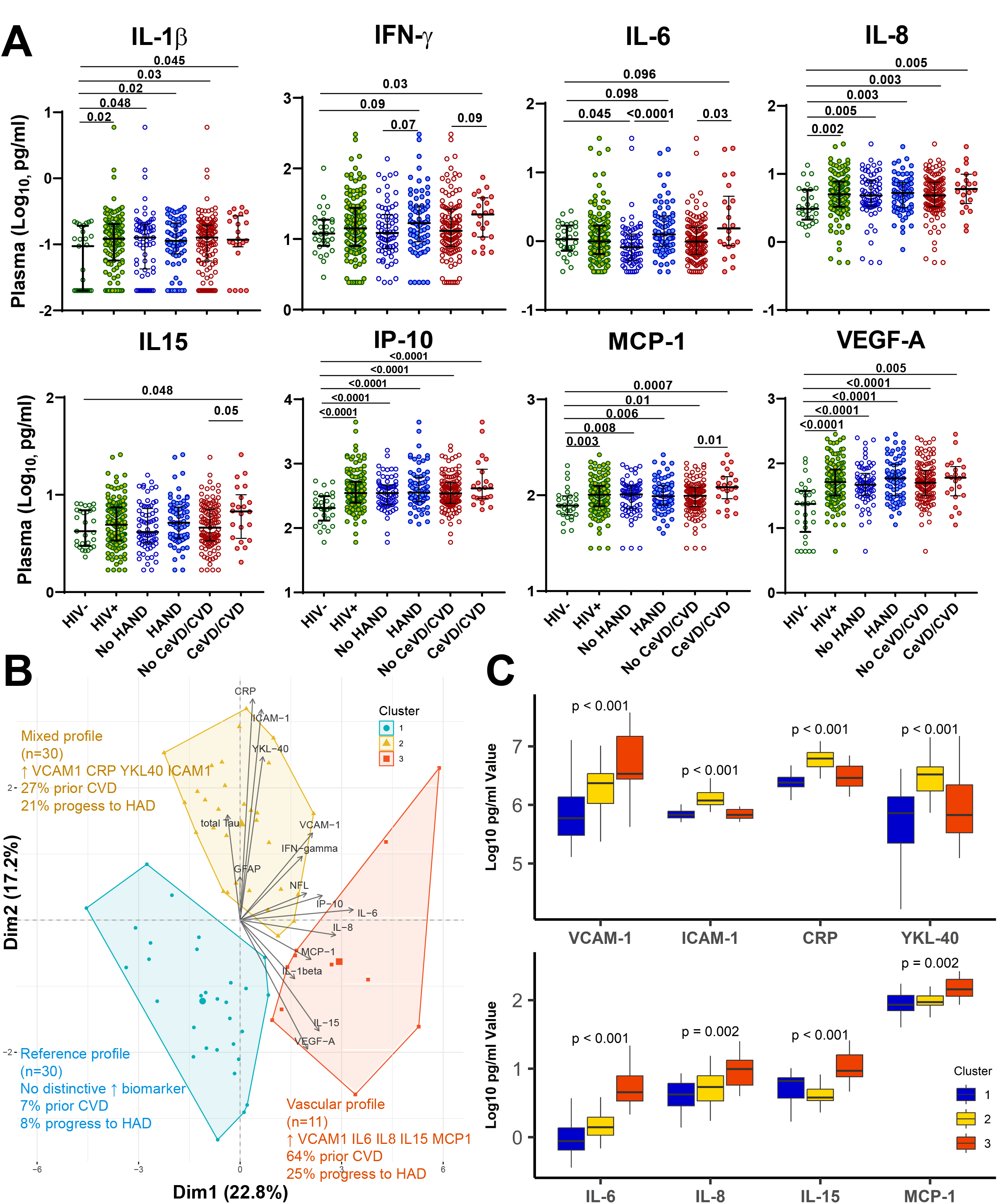
Association of plasma inflammation markers with HIV infection, HAND, and vascular disease. (A) Association of plasma IL-1β, IFN-γ, IL-6, IL-8, IL-15, IP-10, MCP-1, and VEGF-A with HIV infection, HAND, and vascular disease. Medians and IQRs are indicated as horizontal and vertical lines, respectively. Statistical significance was calculated using Mann– Whitney U test; significant differences (p<0.05) are indicated. (B) and (C) K-means clustering of 15 plasma biomarkers combined with dimensionality reduction by principal component analysis (PCA) among PWH with HAND (n=71) identifies three clusters distinguished by inflammation marker profiles, prevalence of prior vascular disease, and progression to HAD within 2.5 years. Contributions of each biomarker to Dim1 and Dim2 are shown on the PCA plot (B) and Supplementary Digital Content 7. Statistical significance was calculated using the Kruskal-Wallis H test.

### Biomarker clustering separates HAND into clusters distinguished by inflammation profiles, prevalence of vascular disease, and progression to HAD

K-means clustering based on 15 plasma biomarkers in 71 individuals with HAND combined with dimensionality reduction by PCA identified three distinct clusters (Figure 3B). Among 71 individuals with HAND, 62 had 1 or more follow-up visits with neurocognitive evaluations at 6- or 12-month intervals including 10 (16%) who progressed to HAD within 2.5 years. The 1^st^ and 2^nd^ principal components (Dim1 and Dim2) explained 22.8% and 17.2% of the variance, respectively, mainly driven by IL-6, IL-8, IP-10, VCAM-1, MCP-1, VEGF-1, and NFL (Dim1) and CRP, ICAM-1, YKL-40, VEGF-A, and IL-15 (Dim2) (Figure 3B and Supplementary Digital Content 7). Biomarker and clinical characteristics according to each cluster are shown in Figure 3C and Supplementary Digital Content 8. Cluster 1 (n=30) represented a reference profile with no distinctive biomarker elevations, younger age (13% over age 60), and relatively lower rates of hypertension (37%), hyperlipidemia (23%), vascular disease (7%), and progression to HAD within 2.5 years (8%). Cluster 2 (n=30) represented a mixed profile characterized by increased plasma VCAM-1, NFL, total Tau, CRP, ICAM-1, and YKL-40, more individuals over age 60 (20%), and relatively higher rates of hypertension (47%), vascular disease (27%) and progression to HAD within 2.5 years (21%). Cluster 3 (n=11) represented a vascular profile characterized by increased plasma VCAM-1, NFL, IL-6, IL-8, IL-15, and MCP-1, high proportion of individuals over age 60 (46%), and highest rates of hypertension (82%), hyperlipidemia (64%), vascular disease (64%) and progression to HAD within 2.5 years (25%). Regarding other distinctive features, cluster 3 had more current smokers compared with clusters 1 and 2 (73% vs. 50% and 57%, respectively), while cluster 2 had lower global T scores compared with clusters 1 and 3 (median 40.9 vs. 43.6 and 43.5, respectively; p=0.043).

## Discussion

In this study, vascular injury markers VCAM-1, ICAM-1, and CRP were associated with HAND and lower global neurocognitive T scores in PWH on suppressive ART. Furthermore, VCAM-1 correlated with the neuronal injury marker NFL. Consistent with these findings, cardiovascular risk factors and diagnoses were more prevalent in HAND vs. no HAND and biomarker clustering identified HAND subgroups distinguished by high prevalence of vascular disease and progression to HAD. In the current ART era, HAND frequently has non-HIV-etiologies, with vascular disease being one of the most common [10, 19–21, 46], and neurocognitive profiles can resemble VCI [10, 17, 19–21, 24, 25, 46]. Although plasma inflammation markers (e.g., IL-1β, IL-8, IP-10, MCP-1) were increased in PWH, only IL-6 was associated with HAND (particularly MND). These findings suggest that vascular injury markers are a stronger predictor of cognitive impairment and CNS injury in virally suppressed PWH than inflammation markers and may help to distinguish relative contributions of VCI to HAND.

Markers of endothelial dysfunction, including VCAM-1 and ICAM-1, are predictors of severity and progression of vascular disease and have been associated with VCI in the general population [47]. Furthermore, increased VCAM-1 was associated with deficits in executive function, processing speed, and working memory in a recent study of 84 PWH [25]. VCAM-1 and ICAM-1 are adhesion molecules that accelerate vascular inflammation and atherosclerosis by increasing attachment of circulating leukocytes to endothelial cells, while CRP is a marker of systemic inflammatory responses that promote vascular disease [18]. Our finding that ICAM-1 and CRP were increased in ART-treated PWH vs. HIV-controls is consistent with previous reports [27, 28, 32]. In contrast to some previous studies [25, 26], we did not detect a significant increase in VCAM-1 in ART-treated PWH vs. HIV-controls. Nonetheless, VCAM-1 and ICAM-1 were associated with HAND, VCAM-1 was higher in MND compared with ANI, and VCAM-1, ICAM-1, and CRP were associated with lower global T scores in age-adjusted regression models. Moreover, VCAM-1 correlated positively with NFL, suggesting association with ongoing CNS injury. Accordingly, VCAM-1 is a potential biomarker for elevated risk of VCI in PWH and may be useful for monitoring responses to interventions targeting vascular disease.

IL-6, CRP, and MCP-1 are inflammation markers associated with vascular disease in PWH [30, 32, 48]. Despite marked reduction of inflammation in PWH on ART, low-level inflammation persists and contributes to vascular and neurological comorbidities [5, 8, 18, 45]. Our finding that PWH with vascular disease had higher plasma IL-6 and MCP-1 compared to those without vascular disease is consistent with previous studies [30, 32, 49] and role of inflammation in promoting atherosclerosis [15, 18, 27, 31]. Furthermore, biomarker clustering separated HAND into three clusters: two clusters with high prevalence of vascular disease, elevated VCAM-1 and NFL, and distinctive inflammation profiles (CRP/ICAM-1/YKL-40 or IL-6/IL-8/IL-15/MCP-1) and one cluster with no distinctive biomarker elevations, which may represent legacy effects of untreated HIV and its complications [3]. Together, these findings raise the possibility that targeting vascular inflammation may be beneficial to prevent or ameliorate VCI in PWH [48].

YKL-40, a biomarker of cardiovascular disease and neuroinflammation [50–52], correlated positively with vascular injury markers and was increased in plasma and CSF of PWH with HAND vs. no HAND. YKL-40 can be generated from multiple sources including macrophages/microglia, astrocytes, chondrocytes, neutrophils, and fibrotic liver tissue, and is involved in peripheral and CNS inflammation, tissue remodeling, and vascular disease [50, 52]. While the role of CSF YKL-40 as a prognostic marker for HAND [34, 39] and Alzheimer’s disease [51] has been documented, the role of plasma YKL-40 in relation to cognitive impairment in PWH is not well studied and warrants further evaluation to understand its relationship to biologically defined subtypes of HAND.

Plasma NFL is a sensitive biomarker of CNS injury in HAND and other neurological disorders [33, 38, 53–55]. In our study, plasma NFL was increased in PWH compared with uninfected controls, albeit not significantly different between HAND vs. no HAND. Plasma NFL correlated negatively with global T scores and the association remained significant in models adjusted for age, suggesting an association with subclinical CNS injury. Moreover, vascular disease was associated with increased plasma NFL. Previous studies detected increased plasma or CSF NFL in people with cerebral small vessel disease-related white matter injury [56, 57], a common pathology in PWH [21, 24, 46] and correlate of VCI in the general population [47]. Total Tau, a biomarker of neurodegeneration in Alzheimer’s disease and other neurodegenerative disorders, showed an increasing trend in CSF, but not plasma, in HAND compared with no HAND. Previous studies of CSF total Tau in PWH reported inconsistent findings, which may reflect differences in study design such as cohort age, viral suppression, and severity of cognitive impairment [12, 35, 36, 58]. Further studies are warranted to understand the prognostic value of NFL, Tau, and other markers of CNS injury in older PWH in relation to current and future risk of cognitive impairment.

We acknowledge several limitations of the study. Our study was limited by the sample size and fewer participants with available CSF samples, which limited statistical power to detect some associations. Although we detected a modest increase of CSF NFL in cognitively impaired PWH in a previous study that included some participants with HAD or viremia [59], similar to findings reported by others [33, 38, 45, 55], we did not detect an association of HAND with CSF NFL in the present study. Other limitations are the low proportion of PWH on integrase inhibitors (19%) and newer ART drugs, and lack of available neuroimaging data. Additionally, we did not evaluate the relationship of increased VCAM-1 to deficits in specific cognitive domains [25]. Lastly, as this was a cross-sectional study, we could not evaluate prospective associations of biomarkers with future development of HAND. Longitudinal studies of larger cohorts including older participants and newer ART drugs are needed to overcome these limitations and identify biomarkers associated with progression of vascular injury and HAND in ART-treated PWH.

In summary, we present evidence that peripheral markers of vascular injury are more closely associated with HAND and CNS injury in PWH on suppressive ART than markers of inflammation. Our findings are consistent with the three-hit model proposed by Jakabek et al [20] in which HIV infection, aging, and vascular disease contribute to brain aging in PWH, and suggest that vascular injury markers, particularly VCAM-1, may help to distinguish relative contributions of VCI to HAND. Further studies are warranted to better understand the utility of VCAM-1 and other peripheral markers of vascular injury for identification of HAND biotypes [11, 13] and development of tailored interventions in virally suppressed PWH on ART.

## Data Availability

All data generated in the study are included in the published article and its Supplementary Information files or available from the corresponding author upon reasonable request.

## Author contributions

DG (first author) participated in study design, performed experiments, data assembly, statistical analysis, and interpretation, wrote the initial manuscript draft, and prepared tables and figures. VM participated in performing experiments, data assembly, and manuscript editing. JY, MH, and HU performed statistical analyses and prepared tables and figures. DG (last author) designed and supervised the study, coordinated assembly and organization of data, participated in data analysis and interpretation, and drafting and editing the manuscript, figures, and tables. All authors read, participated in editing the manuscript, and approved the final manuscript.

## Funding

Supported by NIH grants to D.G (R01MH110259 and R01DA046203). NNTC sites were supported by National Institute of Mental Health (NIMH) and National Institute of Neurological Disorders and Stroke (NINDS) (grants U24MH100931, U24MH100930, U24MH100929, U24MH100928, U24MH100925). CHARTER sites were supported by HHSN271201000036C and HHSN271201000030C from NIMH/NINDS.

## List of Supplemental Digital Content

**Supplementary Digital Content 1.**
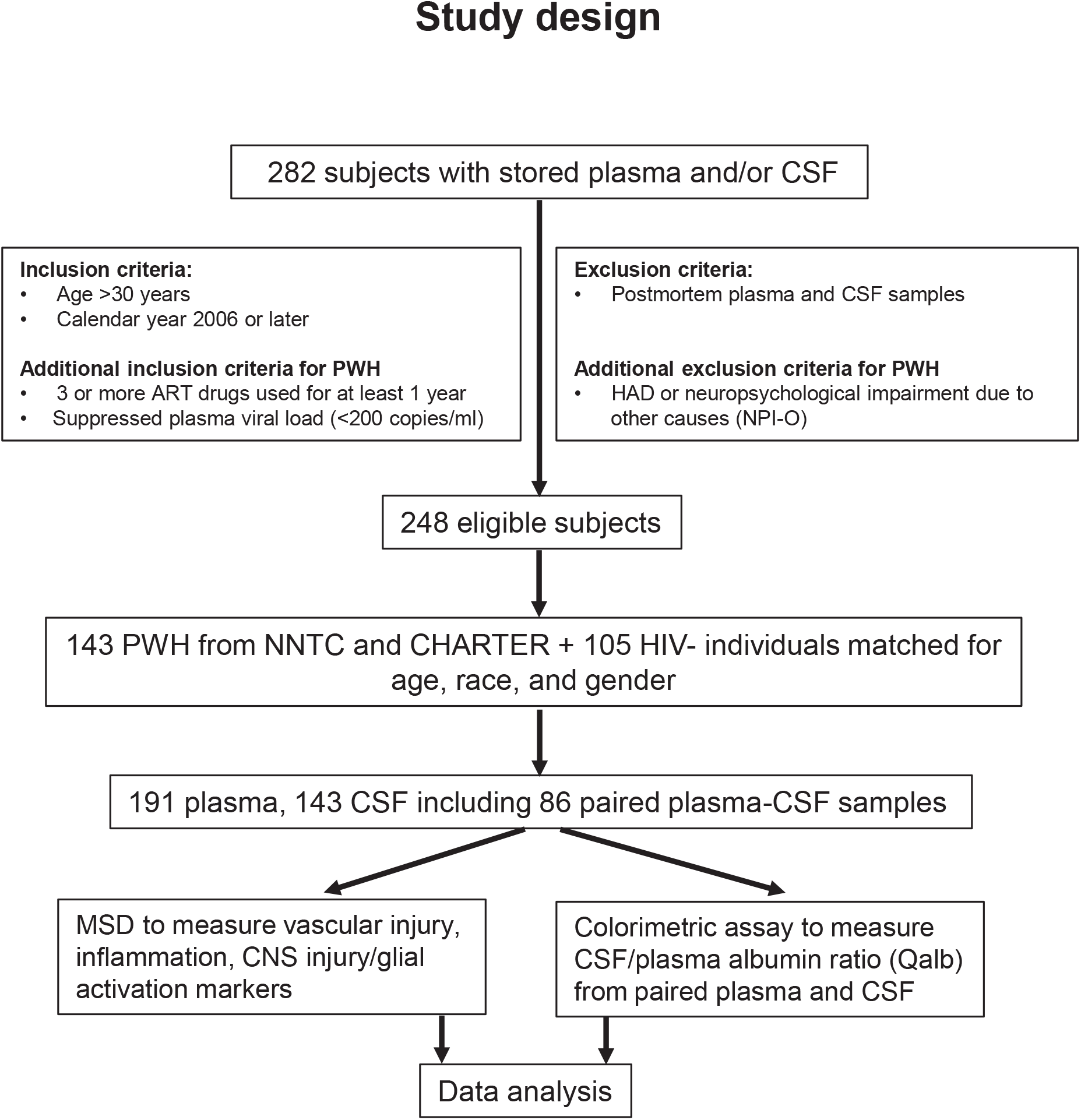
Overview of cohort selection and study design. From a sample of 282 individuals with stored plasma and/or CSF samples, we identified 248 eligible individuals meeting inclusion criteria and with sufficient volume of sample to perform biomarker assays.

**Supplementary Digital Content 2.**
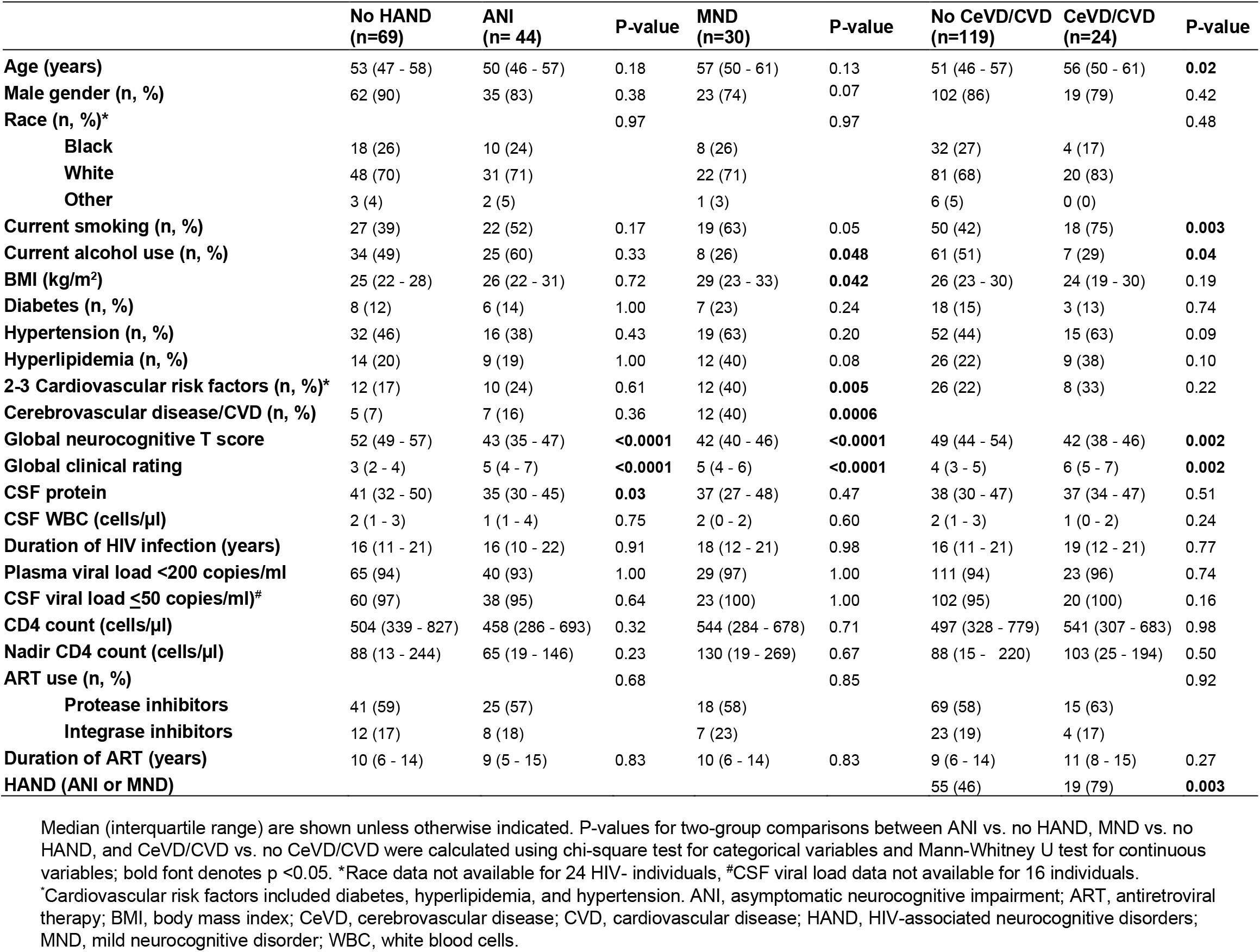
Demographic and clinical characteristics of study participants by HAND and vascular disease status.

**Supplementary Digital Content 3.**
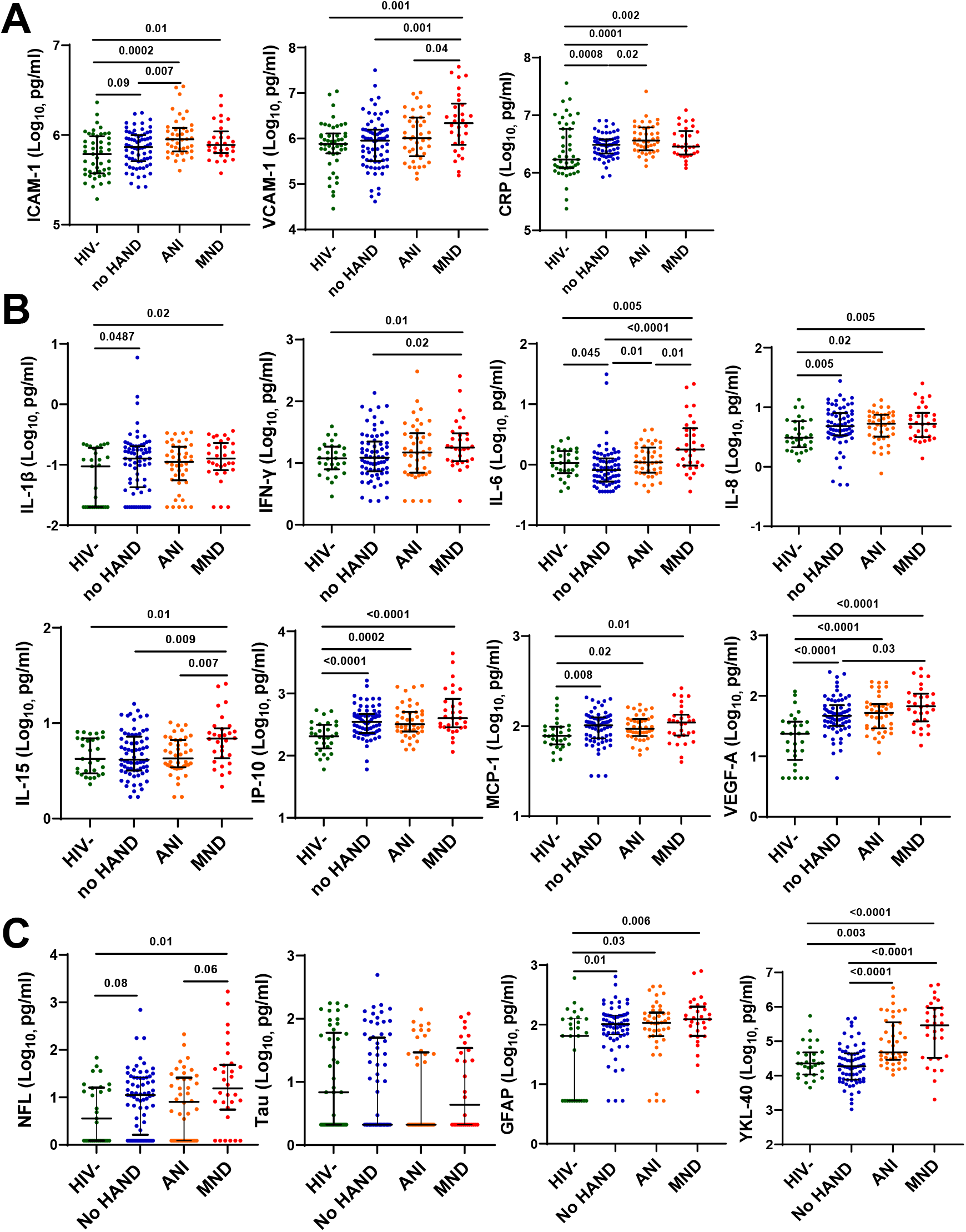
Association of plasma vascular injury, inflammation, and CNS injury markers with ANI or MND. (A) Association of plasma ICAM-1, VCAM-1, and CRP, (B) IL-1β, IFN-γ, IL-6, IL-8, IL-15, IP-10, MCP-1, and VEGF-A, and (C) NFL, Tau, GFAP, and YKL-40 with ANI or MND vs. no HAND or HIV-control groups. Medians and IQRs are indicated as horizontal and vertical lines, respectively. Statistical significance was calculated using Mann–Whitney U test; significant differences (p<0.05) are indicated.

**Supplementary Digital Content 4.** Association of plasma NFL, GFAP, YKL-40, ICAM-1, VCAM-1, and CRP with global neurocognitive T scores < 40 in logistic regression models adjusted for age.

| **Variable** | **$\beta$** | **SE** | **95% CI** | **P-value** |
| --- | --- | --- | --- | --- |
| **Intercept** | 0.84 | 1.64 | -2.40 to 4.11 | 0.61 |
| **Age** | -0.070 | 0.034 | -0.14 to -0.005 | 0.037 |
| **NFL** | 1.057 | 0.49 | 0.11 to 2.06 | 0.030 |
| **Intercept** | -3.46 | 1.83 | -7.19 to 0.039 | 0.059 |
| **Age** | -0.014 | 0.026 | -0.066 to 0.038 | 0.59 |
| **GFAP** | 1.29 | 0.76 | -0.14 to 2.85 | 0.090 |
| **Intercept** | -9.40 | 3.002 | -15.73 to -3.82 | 0.002 |
| **Age** | 0.006 | 0.026 | -0.044 to 0.057 | 0.82 |
| **YKL-40** | 1.23 | 0.42 | 0.45 to 2.12 | 0.004 |
| **Intercept** | -16.31 | 6.83 | -30.25 to -3.16 | 0.017 |
| **Age** | 0.011 | 0.026 | -0.040 to 0.064 | 0.67 |
| **ICAM-1** | 2.38 | 1.084 | 0.29 to 4.58 | 0.028 |
| **Intercept** | -8.78 | 2.79 | -14.55 to -3.53 | 0.002 |
| **Age** | -0.006 | 0.026 | -0.058 to 0.045 | 0.81 |
| **VCAM-1** | 1.22 | 0.41 | 0.45 to 2.060 | 0.003 |
| **Intercept** | -11.74 | 6.61 | -25.02 to 1.13 | 0.076 |
| **Age** | 0.007 | 0.026 | -0.044 to 0.058 | 0.80 |
| **CRP** | 1.50 | 0.96 | -0.38 to 3.42 | 0.12 |

Multivariable logistic regression models were fit for plasma NFL, GFAP, YKL-40, ICAM-1, VCAM-1, and CRP in PWH with global neurocognitive T scores <40 vs. ≥ 40 as the dependent variable adjusting for age. Biomarker variables were log<sub>10</sub> transformed. Abbreviations: SE, standard error; CI, confidence interval.

**Supplementary Digital Content 5.**
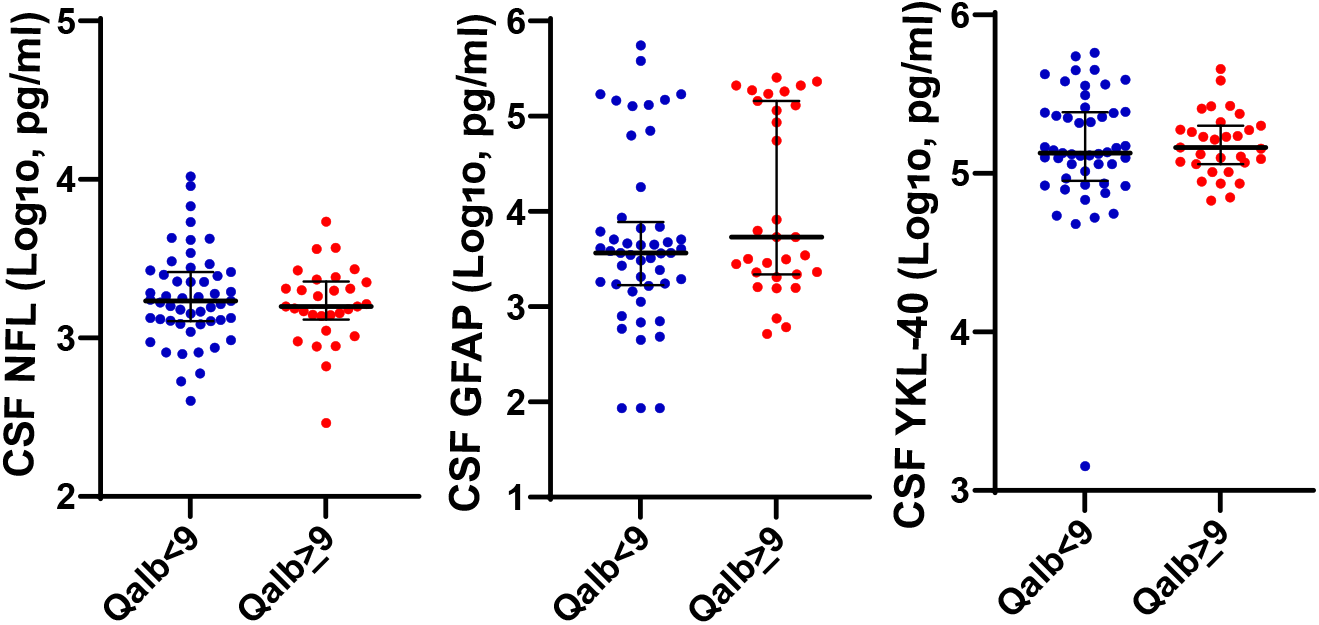
CSF markers of CNS injury have no significant association with CSF/plasma albumin ratio (Qalb). Abnormally increased Qalb (values >9, corresponding to increased BBB permeability vs. <9, considered as normal) is not associated with CSF NFL, GFAP, and YKL-40 levels. Medians and IQRs are indicated as horizontal and vertical lines, respectively. Statistical significance was calculated using Mann– Whitney U test; significant differences (p<0.05) are indicated.

**Supplementary Digital Content 6.**
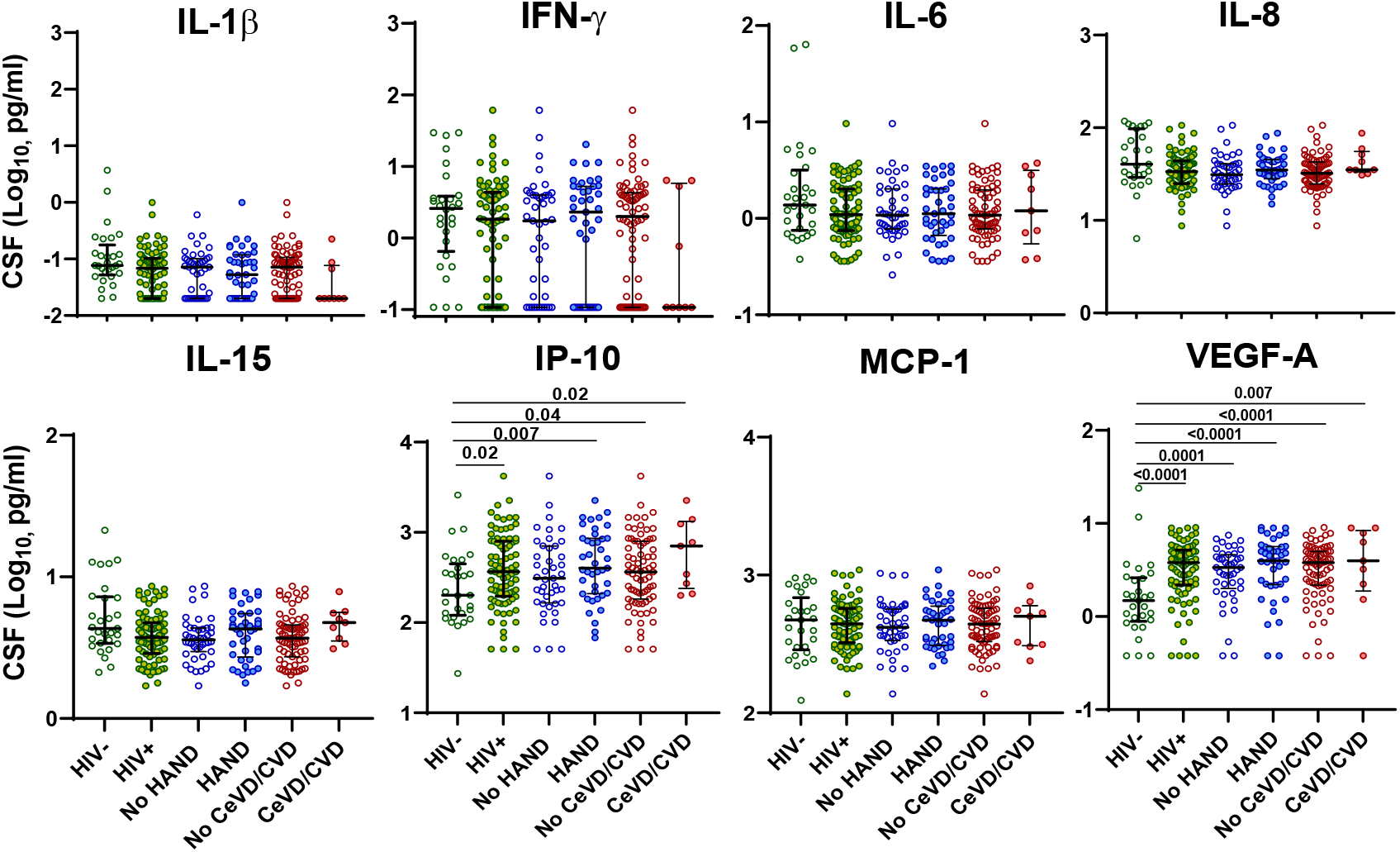
CSF inflammation markers IL-1β, IFN-γ, IL-6, IL-8, IL-15, IP-10, MCP-1, and VEGF have no significant association with HAND or vascular disease. Medians and IQRs are indicated as horizontal and vertical lines, respectively. Statistical significance was calculated using Mann–Whitney U test; significant differences (p<0.05) are indicated.

**Supplementary Digital Content 7.**
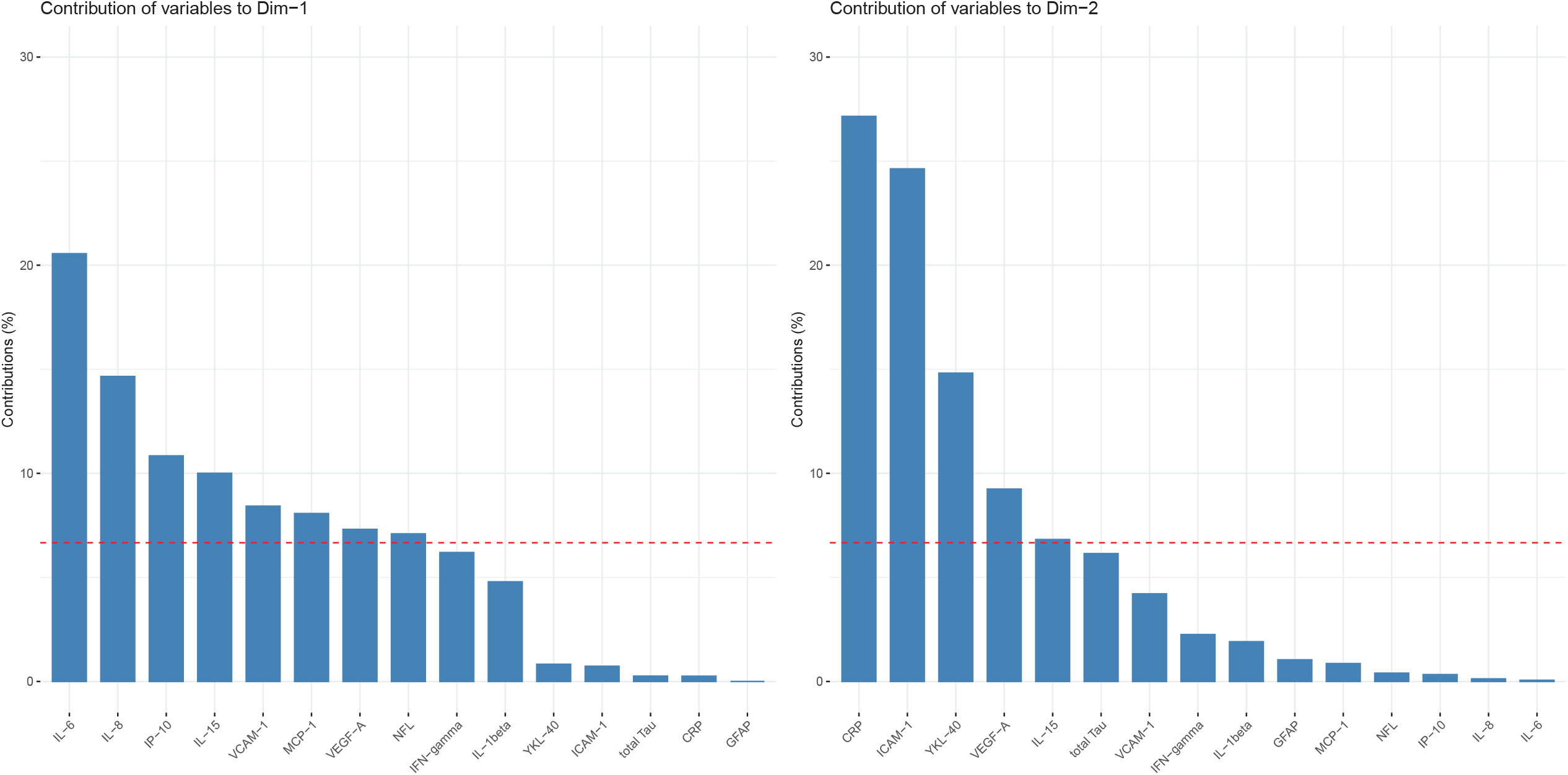
Contribution of each of the 15 variables to the 1st and 2nd principal components (Dim1 and Dim2). Bar charts show the contribution of each variable (%) to Dim1 and Dim2. The total sum of contributions is 100% for each principal component. The red dashed line indicates the expected average contribution of each variable, i.e., 100%/15 = 6.6%.

**Supplementary Digital Content 8.** Association of plasma biomarker clusters with clinical characteristics among HIV+ individuals with HAND.

| Variable |  | Cluster 1<br>(n=30) | Cluster 2<br>(n=30) | Cluster 3<br>(n=11) | P-value |
| --- | --- | --- | --- | --- | --- |
| IL-1β (Log<sub>10</sub>, pg/ml) | Median (Q1, Q3) | -0.9 (-1.1, -0.8) | -1.0 (-1.2, -0.7) | -0.7 (-1.0, -0.5) | 0.190 |
| IFN-γ (Log<sub>10</sub>, pg/ml) | Median (Q1, Q3) | 1.2 (0.9, 1.3) | 1.2 (0.9, 1.6) | 1.5 (1.2, 1.7) | **0.047** |
| IL-6 (Log<sub>10</sub>, pg/ml) | Median (Q1, Q3) | -0.1 (-0.2, 0.1) | 0.1 (0.0, 0.3) | 0.7 (0.5, 0.9) | **<0.001** |
| IL-8 (Log<sub>10</sub>, pg/ml) | Median (Q1, Q3) | 0.6 (0.5, 0.8) | 0.7 (0.5, 0.9) | 1.0 (0.8, 1.1) | **0.002** |
| IL-15 (Log<sub>10</sub>, pg/ml) | Median (Q1, Q3) | 0.8 (0.6, 0.9) | 0.6 (0.5, 0.7) | 1.0 (0.9, 1.2) | **<0.001** |
| IP-10 (Log<sub>10</sub>, pg/ml) | Median (Q1, Q3) | 2.5 (2.4, 2.6) | 2.6 (2.5, 2.8) | 2.9 (2.6, 3.2) | **0.010** |
| MCP-1 (Log<sub>10</sub>, pg/ml) | Median (Q1, Q3) | 1.9 (1.8, 2.1) | 2.0 (1.9, 2.1) | 2.2 (2.1, 2.3) | **0.002** |
| VEGF-A (Log<sub>10</sub>, pg/ml) | Median (Q1, Q3) | 1.8 (1.6, 2.0) | 1.7 (1.4, 1.9) | 1.8 (1.7, 2.0) | 0.124 |
| ICAM-1 (Log<sub>10</sub>, pg/ml) | Median (Q1, Q3) | 5.8 (5.8, 5.9) | 6.1 (6.0, 6.2) | 5.8 (5.8, 5.9) | **<0.001** |
| VCAM-1 (Log<sub>10</sub>, pg/ml) | Median (Q1, Q3) | 5.8 (5.5, 6.1) | 6.4 (6.0, 6.5) | 6.5 (6.4, 7.2) | **<0.001** |
| CRP (Log<sub>10</sub>, pg/ml) | Median (Q1, Q3) | 6.4 (6.3, 6.5) | 6.8 (6.7, 6.9) | 6.5 (6.3, 6.7) | **<0.001** |
| NFL (Log<sub>10</sub>, pg/ml) | Median (Q1, Q3) | 0.4 (0.1, 1.1) | 1.2 (0.6, 1.6) | 1.7 (1.5, 2.4) | **<0.001** |
| Total Tau (Log<sub>10</sub>, pg/ml) | Median (Q1, Q3) | 0.3 (0.3, 1.3) | 1.3 (0.3, 1.8) | 0.3 (0.3, 1.2) | 0.079 |
| GFAP (Log<sub>10</sub>, pg/ml) | Median (Q1, Q3) | 1.9 (1.7, 2.2) | 2.0 (1.8, 2.2) | 2.2 (2.1, 2.4) | **0.045** |
| YKL-40 (Log<sub>10</sub>, pg/ml) | Median (Q1, Q3) | 5.9 (5.3, 6.1) | 6.5 (6.2, 6.6) | 5.8 (5.5, 6.3) | **<0.001** |
| Age (60 years or older) | N (%) | 4 (13.3) | 6 (20.0) | 5 (45.5) | 0.085 |
| Female gender | N (%) | 7 (23.3) | 5 (16.7) | 3 (27.3) | 0.791 |
| Race | Black, N (%) | 7 (23.3) | 7 (23.3) | 4 (36.4) | 0.858 |
|  | White, N (%) | 21 (70.0) | 22 (73.3) | 7 (63.6) |  |
|  | Other, N (%) | 2 (6.7) | 1 (3.3) | 0 |  |
| Current smoking | N (%) | 15 (50.0) | 17 (56.7) | 8 (72.7) | 0.473 |
| Current alcohol use | N (%) | 18 (60.0) | 12 (40.0) | 2 (18.2) | 0.051 |
| BMI | Median (Q1, Q3) | 27.3 | 26.9 | 24.1 | 0.506 |
|  |  | (23.5, 31.6) | (21.7, 33.0) | (20.7, 28.7) |  |
| Diabetes | N (%) | 6 (20.0) | 3 (10.0) | 3 (27.3) | 0.360 |
| Hypertension | N (%) | 11 (36.7) | 14 (46.7) | 9 (81.8) | **0.043** |
| Hyperlipidemia | N (%) | 7 (23.3) | 6 (20.0) | 7 (63.6) | **0.025** |
| 2-3 Cardiovascular risk factors* | N (%) | 9 (30.0) | 5 (16.7) | 7 (63.6) | **0.014** |
| Cerebrovascular disease/CVD | N (%) | 2 (6.7) | 8 (26.7) | 7 (63.6) | **0.001** |
| Global neurocognitive T score | Median (Q1, Q3) | 43.6 | 40.9 | 43.5 | **0.043** |
|  |  | (40.1, 49.8) | (35.2, 43.9) | (40.0, 48.4) |  |
| Global clinical rating | Median (Q1, Q3) | 5.0 (4.0, 6.0) | 5.0 (5.0, 7.0) | 5.0 (4.2, 5.8) | 0.055 |
| CSF protein | Median (Q1, Q3) | 39.0 | 34.7 | 34.0 | 0.194 |
|  |  | (32.0, 47.0) | (29.5, 42.3) | (26.9, 37.0) |  |
| CSF WBC | Median (Q1, Q3) | 1.0 (0.5, 4.0) | 1.0 (1.0, 2.0) | 2.0 (1.2, 2.0) | 0.844 |
| Duration of HIV infection (years) | Median (Q1, Q3) | 18.3 | 15.1 | 16.2 | 0.962 |
|  |  | (12.2, 21.4) | (10.5, 22.2) | (12.6, 19.3) |  |
| Plasma viral load <200 copies/ml | N (%) | 27 (93.1) | 28 (93.3) | 11 (100) | 1.000 |
| CSF viral load ≤50 copies/ml<sup>#</sup> | N (%) | 24 (88.9) | 29 (100) | 6 (100) | 0.220 |
| CD4 count (cells/μl) | Median (Q1, Q3) | 523.5 | 495.5 | 627.0 | 0.480 |
|  |  | (309.2, 691.5) | (291.2, 648.8) | (434.0, 841.0) |  |
| Nadir CD4 count (cells/μl) | Median (Q1, Q3) | 45.5 | 95.0 | 133.0 | 0.396 |
|  |  | (16.2, 215.0) | (21.8, 144.2) | (77.0, 253.5) |  |
| Duration of ART (years) | Median (Q1, Q3) | 10.4 (5.2, 14.8) | 8.5 (6.6, 11.0) | 13.0 (9.8, 16.9) | 0.145 |
| Progression to HAD within 2.5 years | N (%) | 2 (8.0) | 6 (20.7) | 2 (25.0) | 0.311 |

P-values for comparisons between groups were calculated using Fisher's exact test for categorical variables and Kruskal-Wallis H test for continuous variables; bold font denotes $p < 0.05$ . \*2-3 cardiovascular risk factors included diabetes, hyperlipidemia, and hypertension. <sup>#</sup>CSF viral load data not available for 9 individuals. ART, antiretroviral therapy; BMI, body mass index; CVD, cardiovascular disease; HAD, HIV-associated dementia; HAND, HIV-associated neurocognitive disorders; WBC, white blood cells.

## Notes

### Competing Interest Statement

The authors have declared no competing interest.

### Funding Statement

The study was funded by NIH grants to D.G (R01MH110259 and R01DA046203). NNTC sites were supported by National Institute of Mental Health (NIMH) and National Institute of Neurological Disorders and Stroke (NINDS) (grants U24MH100931, U24MH100930, U24MH100929, U24MH100928, U24MH100925). CHARTER sites were supported by HHSN271201000036C and HHSN271201000030C from NIMH/NINDS.

### Author Declarations

The study was approved by the Institutional Review Board of Dana Farber Cancer Institute (DFCI protocol 16-273) and was considered exempt due to using only de-identified data. The NNTC and CHARTER cohorts which provided samples and data for the study were approved by the Institutional Review Boards of all the participating institutions (University of Texas Medical Branch Galveston, University of California Los Angeles, Icahn School of Medicine at Mount Sinai, University of California San Diego, Johns Hopkins University, University of Washington, and Washington University).

